# HIBRID: Histology and ct-DNA based Risk-stratification with Deep Learning

**DOI:** 10.1101/2024.07.23.24310822

**Authors:** Chiara M.L. Loeffler, Hideaki Bando, Srividhya Sainath, Hannah Sophie Muti, Xiaofeng Jiang, Marko van Treeck, Nic Gabriel Reitsam, Zunamys I. Carrero, Tomomi Nishikawa, Toshihiro Misumi, Saori Mishima, Daisuke Kotani, Hiroya Taniguchi, Ichiro Takemasa, Takeshi Kato, Eiji Oki, Tanwei Yuan, Durgesh Wankhede, Sebastian Foersch, Hermann Brenner, Michael Hoffmeister, Yoshiaki Nakamura, Takayuki Yoshino, Jakob Nikolas Kather

**Affiliations:** Else Kroener Fresenius Center for Digital Health, Technical University Dresden, Dresden, Germany; Medical Department 1, University Hospital and Faculty of Medicine Carl Gustav Carus, Technische Universität Dresden, Dresden, Germany; National Center for Tumor Diseases Dresden (NCT/UCC), a partnership between DKFZ, Faculty of Medicine and University Hospital Carl Gustav Carus, TUD Dresden University of Technology, and Helmholtz-Zentrum Dresden - Rossendorf (HZDR), Dresden, Germany; Department of Data Science, National Cancer Center Hospital East, Kashiwa, Japan; Department of Gastroenterology and Gastrointestinal Oncology, National Cancer Center Hospital East, Kashiwa, Japan; Translational Research Support Office, National Cancer Center Hospital East, Kashiwa, Japan; Department for Visceral, Thoracic and Vascular Surgery, University Hospital and Faculty of Medicine Carl Gustav Carus, Technische Universität Dresden, Dresden, Germany; Pathology, Faculty of Medicine, University of Augsburg, Augsburg, Germany; Bavarian Cancer Research Center (BZKF), Augsburg, Germany; Department of Clinical Oncology, Aichi Cancer Center Hospital, Nagoya, Japan; Department of Surgery, Surgical Oncology and Science, Sapporo Medical University, Sapporo, Japan; Department of Surgery, NHO Osaka National Hospital, Osaka, Japan; Department of Surgery and Science, Graduate School of Medical Sciences, Kyushu University, Fukuoka, Japan; Division of Clinical Epidemiology and Aging Research, German Cancer Research Center (DKFZ), Heidelberg, Germany; Institute of Pathology, University Medical Center Mainz, Mainz, Germany; Medical Oncology, National Center for Tumor Diseases (NCT), University Hospital Heidelberg, Heidelberg, Germany

**Keywords:** Deep Learning, molecular residual disease, circulating tumor DNA, colorectal cancer, vision transformers

## Abstract

**Background:** Although surgical resection is the standard therapy for stage II/III colorectal cancer (CRC), recurrence rates exceed 30%. Circulating tumor DNA (ctDNA) emerged as a promising recurrence predictor, detecting molecular residual disease (MRD). However, spatial information about the tumor and its microenvironment is not directly measured by ctDNA. Deep Learning (DL) can predict prognosis directly from routine histopathology slides.

**Methods:** We developed a DL pipeline utilizing vision transformers to predict disease-free survival (DFS) based on histological hematoxylin & eosin (H&E) stained whole slide images (WSIs) from patients with resectable stage II-IV CRC. This model was trained on the DACHS cohort (n=1766) and independently validated on the GALAXY cohort (n=1555). Patients were categorized into high- or low-risk groups based on the DL-prediction scores. In the GALAXY cohort, the DL-scores were combined with the four-weeks post-surgery MRD status measured by ctDNA for prognostic stratification.

**Results:** In GALAXY, the DL-model categorized 307 patients as DL high-risk and 1248 patients as DL low-risk (p<0.001; HR 2.60, CI 95% 2.11-3.21). Combining the DL scores with the MRD status significantly stratified both the MRD-positive group into DL high-risk (n=81) and DL low-risk (n=160) (HR 1.58 (CI 95% 1.17-2.11; p=0.002) and the MRD-negative group into DL high-risk (n=226) and DL low-risk (n=1088) (HR 2.37 CI 95% 1.73-3.23; p<0.001). Moreover, MRD-negative patients had significantly longer DFS when predicted as DL high-risk and treated with ACT (HR 0.48, CI 95% 0.27-0.86; p= 0.01), compared to the MRD-negative patients predicted as DL low-risk (HR=1.14, CI 95% 0.8-1.63; p=0.48).

**Conclusion:** DL-based spatial assessment of tumor histopathology slides significantly improves the risk stratification provided by MRD alone. Combining histologic information with ctDNA yields the most powerful predictor for disease recurrence to date, with the potential to improve follow-up, withhold adjuvant chemotherapy in low-risk patients and escalate adjuvant chemotherapy in high-risk patients.

**Highlights:**

- This study combines MRD status measured by ctDNA with a DL-based risk assessment trained on histological image data to enhance recurrence prediction.
- DL-based spatial assessment of tumor histopathology slides significantly improves the risk stratification provided by MRD alone.
- MRD-negative patients with high DL-based risk had a significantly longer DFS if treated with ACT, compared to MRD-negative and DL low risk patients
- The DL model is fully open-source and publicly available.

## Introduction

Colorectal cancer (CRC) is one of the leading causes of cancer-related deaths worldwide^1^. Surgical resection remains the standard curative therapy in patients with Stage II-III CRC and resectable metastases. Despite advancements in surgical and adjuvant therapies, recurrence rates exceed 30% and 60%^2,3^, respectively. Patients who relapse have an increased mortality risk, hence identifying these patients at an early stage is crucial for optimising follow-up treatment decisions. Current prognostication systems for risk assessment, including imaging techniques, clinicopathological features and molecular data, are moderate predictors for recurrence risk. Similarly, follow-up strategies, such as tumor marker monitoring with carcinoembryonic antigen (CEA), lack sensitivity and specificity in identifying recurrence^4–6^. In particular for stage II CRC, the decision on adjuvant chemotherapy (ACT) is based on diverging risk assessment recommendations provided through international oncological associations^7,8^. Thus, a more fine-grained system for estimating the risk of relapse is required, as no stage-specific survival benefit for adjuvant chemotherapy has been proven. Therefore, new biomarkers for better and more precise prognostication are needed. Circulating tumor DNA (ctDNA) has emerged as a promising minimally invasive biomarker that measures a small fraction of ctDNA in the blood, allowing for the detection of molecular residual disease (MRD) status^9^. Additionally, ctDNA can be used for monitoring treatment response and early prediction of recurrence, as ctDNA positivity after surgery is associated with a higher risk of disease recurrence^10,11^. Previous studies have shown that this correlation had already been found as early as four weeks after primary tumor resection^12^. However, ctDNA analysis alone does not capture the morphological characteristic of the tumor. For instance, information such as histopathological subtype, grading, vascular and lymphatic invasion, as well as the abundance of tumor-infiltrating lymphocytes^13–16^, among many other morphological properties of the tumor microenvironment (TME), have been shown to be prognostically relevant and are reflected in current clinical guidelines^17,18^. Deep Learning (DL) is an artificial intelligence technology which is useful to extract quantitative biomarkers from routinely available clinical data in oncology^19,20^. DL models, trained on histopathological routine hematoxylin and eosin (H&E) tumor slides have been shown to act as survival prediction models outperforming current risk-stratifications systems^21–23^. DL can extract highly relevant information from routine pathology slides of CRC, including presence of microsatellite instability (MSI)^24,25^, gene mutations^25,26^, response to neoadjuvant therapy^27^, and overall survival (OS)^22^. Given the ability of DL to extract meaningful biological information from pathology slides that ctDNA cannot capture, we hypothesise that the combination of MRD assessment with a transformer-based DL risk score from morphology could significantly improve prognosis prediction. In this study, we aim to enhance patient stratification and recurrence prediction in patients with CRC by integrating MRD status derived from ctDNA with a DL-based risk score trained on routine histological images.

## Methods & Materials

### Patient Data Acquisition

In this study, we analysed histological whole slide images (WSIs) of hematoxylin & eosin (H&E) stained tumour tissue of surgically curable CRC from two large cohorts in Germany and Japan (Figure 1A-B, Supplementary Figure 1). The first cohort was the Darmkrebs: Chancen der Verhütung durch Screening Study (DACHS), which includes 1774 WSI’s belonging to 1766 patients and was used as a training cohort (Supplementary Figure 1A). The second cohort was the GALAXY trial from the CIRCULATE-Japan study (UMIN000039205), which includes 1556 WSIs from 1555 patients and was used as an independent external validation cohort (Supplementary Figure 1B). The GALAXY trial comprised ctDNA data measuring the MRD status at the four weeks post-surgery interval: MRD positivity was defined as at least 2 out of 16 tumour-specific ctDNA variants detected above a predefined threshold based on Natera’s method^12,28^. Out of the 1555 patients included in the trial, 241 were MRD-positive and 1314 patients were MRD-negative at the respective 4 weeks interval^12^ (Figure 1B). For both cohorts, disease free survival in months (DFS) was available. DFS marked the time from primary surgery to last follow-up date or last surgery to last follow-up date for patients for which primary surgery date was unavailable.

**Figure 1:**
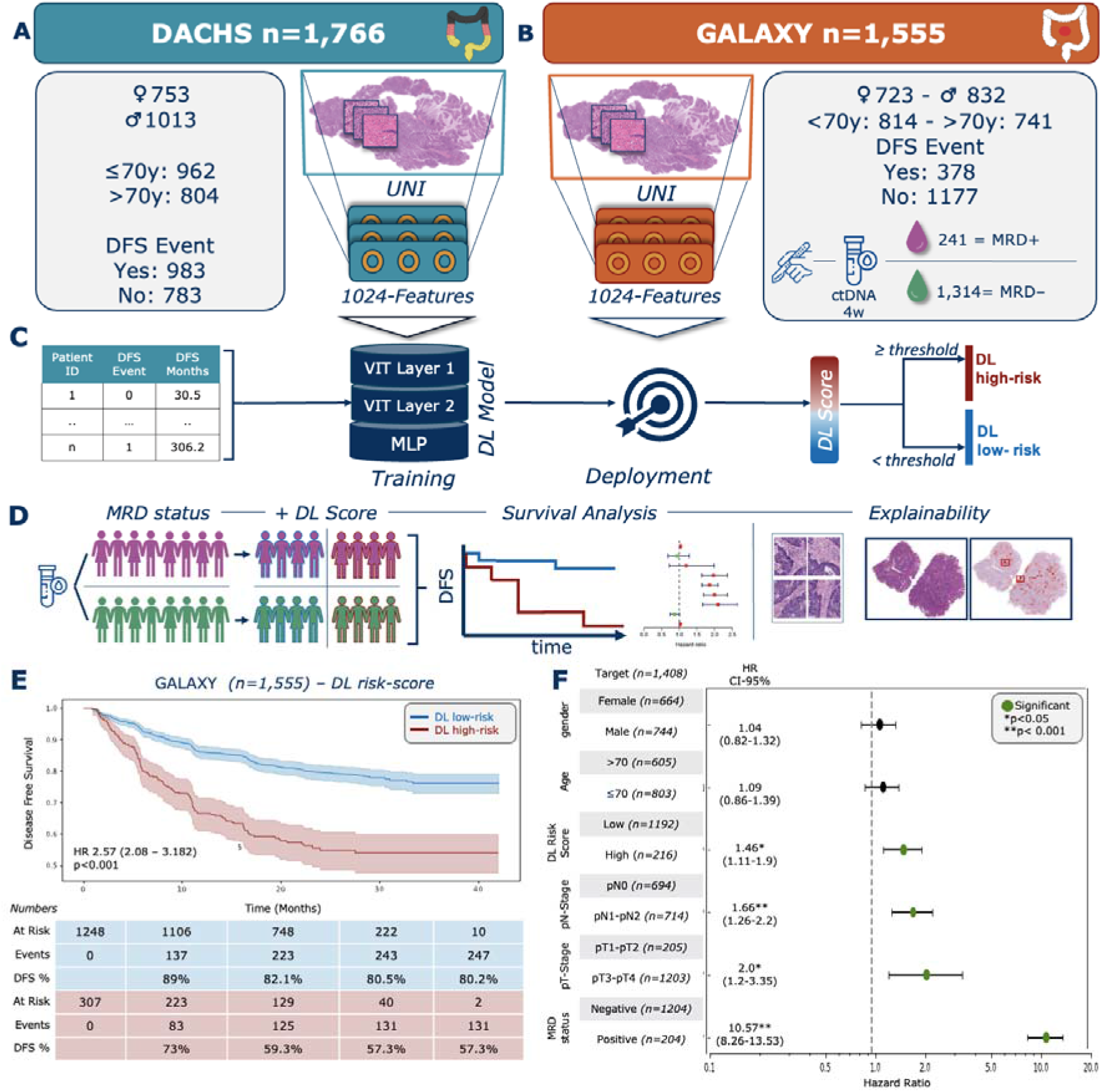
Study Design and DL risk stratification overall. (A) DACHS cohort overview including patient characteristics and WSI preprocessing pipeline using UNI a pretrained vision encoder for feature extraction. (B) GALAXY cohort overview including patient characteristics and WSI preprocessing pipeline. (C) Flowchart of the study design: DFS data was analysed using a Cox-Regression model and fed into the DL-Model combined with the image features from the DACHS cohort for training. The DL-Model was then deployed onto the GALAXY features and a DL-Score was obtained. (D) Overview Experimental Setup: Patients were first categorised based on MRD status and then sub-categorized according to the DL score. Survival analysis with Kaplan-Meier estimator and Cox proportional hazard models were performed. Lastly, highly predictive Tiles and patient whole slide heatmaps were generated. (E) Kaplan-Meier curves for DFS stratified by DL high-risk and DL low-risk patients. (F) Forest plot showing multivariate cox regression analysis including the covariates gender, age, DL risk score, pathological Nodal Stage (pN-Stage), pathological Tumor Stage (pT-Stage) and MRD-status and their association with DFS. HR and 95% CI were calculated by the Cox proportional hazard model. *P*-value was calculated using the two-sided log-rank test (*p<0.05, ** p<0.001). Plot were generated using lifelines package in Python 3.11.5 DACHS=Darmkrebs: Chancen der Verhütung durch Screening Study, WSI=whole-slide image, DFS=disease-free survival, DL=Deep Learning, MRD=molecular residual disease, HR=Hazard ratio, CI=Confidence interval

### Image Processing and Deep Learning Techniques

#### Data Preprocessing

All whole-slide images (WSIs) were segmented into image patches with dimensions of 224□×□224 pixels and an edge length of 256 μm, resulting in an effective magnification of 1·14 μm per pixel. During this segmentation process, patches that primarily contained background or blur (identified by having an average number of Canny edges below a threshold of 2) were removed from the dataset. The retained image tiles were colour normalised using the Macenko method in order to avoid stain-associated bias^29^. For WSI pre-processing, we employed our end-to-end publicly available pipeline, which can be found here: https://github.com/KatherLab/end2end-WSI-preprocessing.

#### Model Development

To train and validate our prediction DL-models we used our open-source pipeline, marugoto (https://github.com/KatherLab/marugoto). In the initial step, a self-supervised learning (SSL) model called UNI, pretrained on over 100 M histology-specific images and 100k WSIs, was employed to extract a 1024-dimensional feature vector from each image tile (Figure 1A-B)^30^. The obtained features were then preprocessed using a multi-headed self-attention mechanism by the transformer network (Figure 1C). Here, the network views patch embeddings as a sequence, with elements interacting through self-attention. For a WSI with n patches of dimension d, self-attention calculates a query-key product. Multi-headed self-attention repeats this in h heads, then concatenating and transforming the outputs. The transformer architecture is designed with two layers, each featuring eight heads (total h=8), a latent dimension of 512, and equal dimensions (each 64) for queries, keys, and values. Post self-attention, the embeddings of each patch are combined into a sequence of dimension n×1024 processed through a linear projection and ReLU activation to reduce dimensionality to 512. A learnable class token is added to this sequence, resulting in an input dimension of (n+1) x 512 that is fed into the transform layer. Each transformer layer consists of a layer normalisation block followed by multi-headed self-attention, a block of layer normalisation and finally a multi-layer perceptron (MLP), with skip connections integrated across each block to facilitate training^24^. After processing through two transformer layers, the class token is inputted into an MLP head designed to produce a continuous risk score for each patient, serving as the output of the model. For model training, we used Cox partial likelihood, as the loss function^22,31^. We randomly split the DACHS cohort at the patient level into training, validation, and test sets in a 4:4:2 ratio. The model was trained using the training set, and the best checkpoint, determined by the highest C-index on the validation set, was saved. This checkpoint was then validated on DACHS test set and CIRCULATE data cohort

#### Visualisation

To interpret our model’s output, we generated whole-slide patient heatmaps showing the DL prediction scores. We used our trained Vision Transformer (ViT) model to process tile-level features extracted from the WSI. The features are passed through the trained model to obtain tile-level scores, which are then combined with Grad-CAM (Gradient-weighted Class Activation Mapping) values to generate weighted scores, which were normalised to a range of −1 to 1 facilitating the identification of the most significant tiles. Heatmaps were then created using the weighted scores, with red indicating high-risk, and blue indicating low-risk. To maintain interpretability, we blended these heatmaps with the original image features, providing clear insights into the tumor morphology and the model’s predictions.

### Experimental Design

In our study we first trained a transformer-based DL model on the DACHS cohort, utilizing clinical data on disease-free survival (DFS) events and DFS time in months to generate patient level DL-based risk scores (Figure 1C). Next, we externally validated the trained DL model on the GALAXY cohort. The continuous DL-risk score was binarized into DL high-risk and DL low-risk categories based on a fixed threshold, defined as the median risk score in the training cohort (0.9357855). Subsequently, we combined the four-week post-surgery MRD status from the GALAXY trial with the DL-risk scores to analyze survival differences between these subgroups (Figure 1D). We also looked at the effects of adjuvant chemotherapy in the various subgroups. Survival Analysis was performed using Kapan-Meier analysis and log-rank test to compare DFS time between the groups. Additionally, multivariate analysis was conducted using Cox proportional hazard models, including the covariates: age, gender, pathological T-Stage (pT) and pathological N-Stage (pN)^22^. Lastly, we performed a morphological analysis to identify histopathological correlations between the DL high-risk and low-risk subgroups, using classification heatmaps (Figure 1D).

### Data and Code availability

Our whole slide image preprocessing pipeline is available here: https://github.com/KatherLab/end2end-WSI-preprocessing. The code for the pretrained vision encoder UNI can be found under: https://github.com/mahmoodlab/uni. Our DL model codes are publicly available at https://github.com/KatherLab/marugoto/tree/survival-transformer/marugoto/survival. The respective study Principal Investigators provided the remaining data. For detailed data sharing policies, please refer to the original publications.

## Results

### DL stratifies patients by recurrence risk

We trained a DL model to generate risk scores based on DFS and validated its performance on the GALAXY cohort. Based on the DL-risk scores, we divided our cohort into DL high- and DL low-risk groups, followed by a survival analysis using Kaplan Meier estimator and Cox proportional hazard analysis (Figure 1D). These results were then compared with the stratification outcomes of MRD status four weeks post-surgery in the GALAXY cohort. Among the 1,555 patients 19.8% (n=307) were categorized as DL high-risk and 80.2% (n=1248) as DL low-risk. Patients classified as DL high-risk exhibited a significantly elevated risk of disease recurrence compared to DL low-risk patients (HR=2.6, CI 95% 2.11-3.21; p < 0.005), with a 20-month DFS of 59.3% vs. 82.1%, respectively (Figure 1D). The ctDNA analysis alone stratified 15.5 % (n=241) patients as MRD-positive and 84.5% (n=1,314) as MRD-negative, with an HR of 11.4 (CI 95% 9.28-14, p<0.001, Supplementary Figure 2A). In the multivariate analysis, including the covariates age, sex, pT, and pN, we found the most prognostic indicator for recurrence risk to be MRD positivity (HR=10.57, CI 95% 8.26-13.53; p<0.001), followed by pT3-pT4-Stage (HR=2.00, CI 95% 1.20-3.35; p<0.05, Figure 1F). The DL-risk score was significant with an HR of 1.46 (CI 95% 1.11-1.90, p<0.05). When correlating the DL risk categories with patient characteristics, we found significant differences in sex, pT-Stage, pN-Stage, pathological Stage, and MRD status (Table 1). Together, these data demonstrate that the DL model can significantly stratify patients according to their risk of recurrence.

**Table 1.** Patients characteristic for DL high-risk and low-risk patients. P values were obtained by a pearsons chi-squared test with Yates’ continuity correction comparing the distribution of the factors between the two columns (DL high-risk vs DL low-risk). Statistical analysis was performed on R 4.4.4. ECOG=Eastern Cooperativ Oncology Group, MSS=microsatellite stable, MSI=microsatellite instable, NA=Not available

| Patient characteristics | Category | DL high-risk (n=307□),<br>n (%) | DL low-risk (n=1248),<br>n (%) | Chi squared and p values |
| --- | --- | --- | --- | --- |
| Age | ≤70 | 167 (54.4) | 647 (51.8) | X<sup>2</sup> = 0.54618<br>P-value = 0.4599 |
|  | >70 | 140 (45.6) | 601 (48.2) |  |
| Sex | Female | 120 (39.1) | 603 (48.3) | X<sup>2</sup> = 8.0696<br>P-value = 0.004501 |
|  | Male | 187 (60.9) | 645 (51.7) |  |
| ECOG performance Status | 0 | 280 (91.2) | 1124 (90) | X<sup>2</sup> = 0.24735<br>P-value = 0.6189 |
|  | 1 | 27 (8.8) | 124 (10) |  |
| pT-Stage | T1-T2 | 10 (3.3) | 195 (15.6) | X<sup>2</sup> = 19.26<br>P-value<0.001 |
|  | T3-T4 | 206 (67.1) | 998 (80) |  |
|  | NA | 91 (29.6) | 55 (4.4) |  |
| pN-Stage | N0 | 76 (35.2) | 618 (51.8) | X<sup>2</sup> = 19.646<br>P-value<0.001 |
|  | N1-2 | 140 (64.8) | 574 (48.2) |  |
|  | NA | 91 (29.6) | 56 (4.5) |  |
| pathological Stage | I | 3 (1) | 148 (11.9) | X<sup>2</sup> = 201.46<br>P-value<0.001 |
|  | II | 65 (21.1) | 452 (36.2) |  |
|  | III | 116 (37.8) | 533 (42.7) |  |
|  | IV | 123 (40.1) | 115 (9.2) |  |
| RAS status | RAS wild-type | 102 (54.8) | 394 () | X<sup>2</sup> = 0.32606<br>P-value=0.568 |
|  | RAS mutant | 84 (45.2) | 291 () |  |
|  | NA | 121 (39.4) | 563 (45.1) |  |
| **BRAF status** | *BRAF* wild-type | 171 (55.7) | 623 (49.9) | $\chi^2 = 1.2724e-28$<br>*P*-value=1 |
|  | *BRAF* mutant | 13 (4.2) | 48 (3.8) |  |
|  | NA | 123 (40.1) | 577 (46.2) |  |
| **MSI status** | MSI | 20 (6.5) | 102 (8.2) | $\chi^2 = 0.46606$<br>*P*-value=0.4948 |
|  | MSS | 262 (85.3) | 1090 (87.3) |  |
|  | NA | 25 (8.1) | 56 (4.5) |  |
| **MRD Status** | MRD-positive | 81 (26.4) | 160 (12.8) | $\chi^2 = 33.585$<br>*P*-value <0.001 |
|  | MRD-negative | 226 (73.6) | 1088 (87.2) |  |

### DL stratifies recurrence risk within MRD subgroups

We hypothesised that by integrating the MRD status with our DL risk score we can further stratify the patients according to risk of recurrence, particularly the MRD-negative patients. To test this, we combined the binarized DL-derived risk score with the MRD status four-weeks after curative surgery (Figure 2). In the MRD-positive group, 33.6% (81 out of 241 patients) were categorized as DL high-risk and 66.4% (160 out of 241 patients) as DL low-risk, with an HR of 1.57 (CI 95% 1.18-2.12; p=0.002, Figure 2A). The DFS-time interval was longer in the DL low-risk group, with a 20-months DFS of 30.9% compared to 9.9% in the DL high-risk group (Figure 2A).

**Figure 2:**
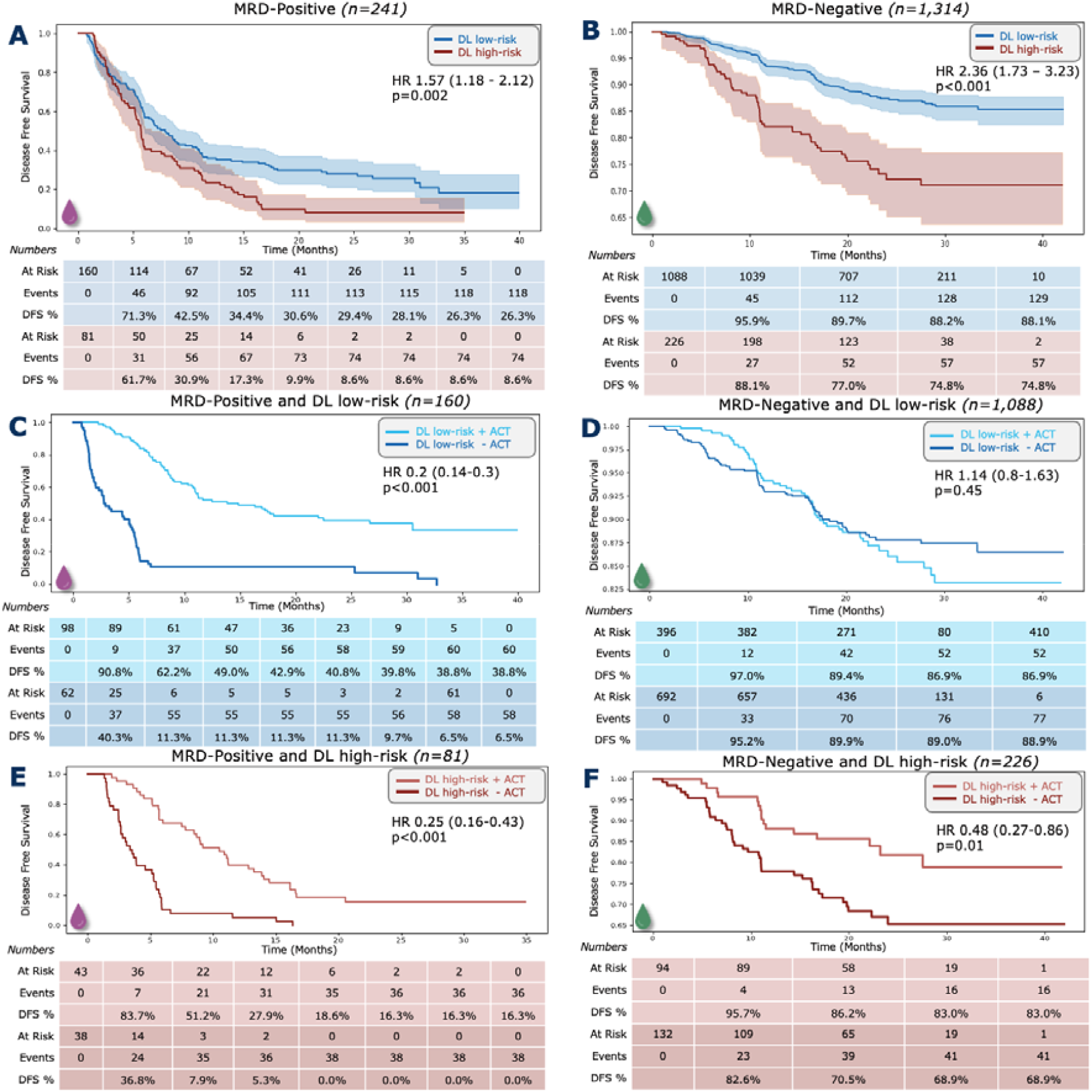
DL stratifies recurrence risk within MRD subgroups. Kaplan-Meier curves showing DFS stratification by DL high-risk and DL low-risk groups for (A) MRD-positive and (B) MRD-negative groups, followed by Kaplan-Meier curves showing DFS stratified by with or without ACT treatment in (C) MRD-positive and DL low-risk, (D) MRD-negative and DL low-risk, (E) MRD-positive and DL high-risk and (F) MRD-negative and DL low-risk subgroups. HR and 95% CI were calculated by the Cox proportional hazard model. *P-v*alue was calculated using the two-sided log-rank test. Plots were generated using the lifelines package in Python 3.11.5 DFS=disease-free survival, DL=Deep Learning, ACT=adjuvant chemotherapy, MRD=molecular residual disease, HR=Hazard ratio, CI=Confidence interval.

In the MRD-negative group, 17.2% (226 out of 1,314 patients) were classified as high-risk by the DL model and 82.8% (1,088 out of 1,314 patients) as DL low-risk with an HR of 2.36 (CI 95% 1.73-3.23; p<0.001, Figure 2B). Additionally, the 20-month DFS was longer in the DL low-risk group at 89.7%, compared to 77% in the DL high-risk group (Figure 2B). In a multivariate Cox analysis with age, sex, pT and pN as covariates, the DL-score was the only independent prognostic predictor in the MRD-positive group with an HR of 1.51 (CI 95% 1.07-2.14; p= 0.018, Supplementary Figure 2B). In the MRD-negative group, pT1-T2 was the strongest prognostic indicator with an HR of 2.50 (CI 95% 1.73-3.64; p<0.001, Supplementary Figure 2C). The DL risk score, with an HR of 1.28 (CI 95% 0.83-1.99; p=0.27), was not an independent prognostic predictor (Supplementary Figure 2C). In summary, these data show that the combination of MRD status with the DL risk score enables a better stratification of patients with CRC.

### DL-based recurrence risk predicts benefit from adjuvant chemotherapy in MRD-negative patients

We hypothesised that our DL-risk score could identify patients with stage II-IV CRC who might benefit from ACT, despite being MRD-negative. To test this hypothesis, we explored the association of ACT with DFS by performing Kaplan-Meier analysis within the DL high-risk and low-risk subgroups among both MRD-positive and MRD-negative patients (Figure 2 C-F). For the MRD-positive group, patients receiving ACT in had significantly longer DFS in both the DL low-risk group (HR=0.20, CI 95% 0.14-0.30; p<0.001, Figure 2C) and in the DL high-risk group (HR=0.25, CI 95% 0.16-0.43; p<0.001, Figure 2E). Without receiving ACT, all MRD-positive and DL high-risk patients experienced recurrence within 20-months, whereas 18.6% of the MRD-positive and DL high-risk patients who received ACT remained disease-free after 20 months (Figure 2E). In the MRD-negative group, patients in the low-risk DL group did not have longer DFS when treated with ACT (HR=1.14, CI 95% 0.8-1.63; p=0.48). The 20-month DFS was 89.4% for patients treated with ACT vs 89.9% for patients not receiving ACT (Figure 2D). Interestingly, patients in the MRD-negative and DL high-risk group showed significantly longer DFS when treated with ACT (HR 0.48, CI 95% 0.27-0.86; p= 0.01, Figure 2D). The 20-month DFS rate was 86.2% in patients who received ACT and thus significantly higher than in patients who did not receive ACT (70.5%). This disease-free survival advantage continued to be seen in the 40-month DFS rate at 83% (with ACT) vs 68.9% (without receiving ACT, Figure 2F).

Together, these data show that the DL prognostication model can successfully further stratify MRD-negative patients. This indicated that even within the low-risk subgroup (according to MRD), there are high-risk individuals for whom the omission of ACT may carry a higher risk of recurrence.

### DL can identify histopathological features linked to prognosis

Measurements of ctDNA provide information about viable and disseminated tumor cells, serving as surrogate markers for their presence in the body and enabling a non-invasive assessment of MRD after surgery. However, they do not provide any information regarding tumor morphology as well as the TME, which is reflected in histopathology slides and is known to be related to clinical outcomes. We investigated whether our model trained on histopathology images without any manual annotation, learned to consider morphological features of the tumor and the TME, which would be synergistic to MRD status. We used a model trained on DACHS and deployed on GALAXY, visualising highly predictive regions at both high and low magnification, as shown in Figure 3.

**Figure 3:**
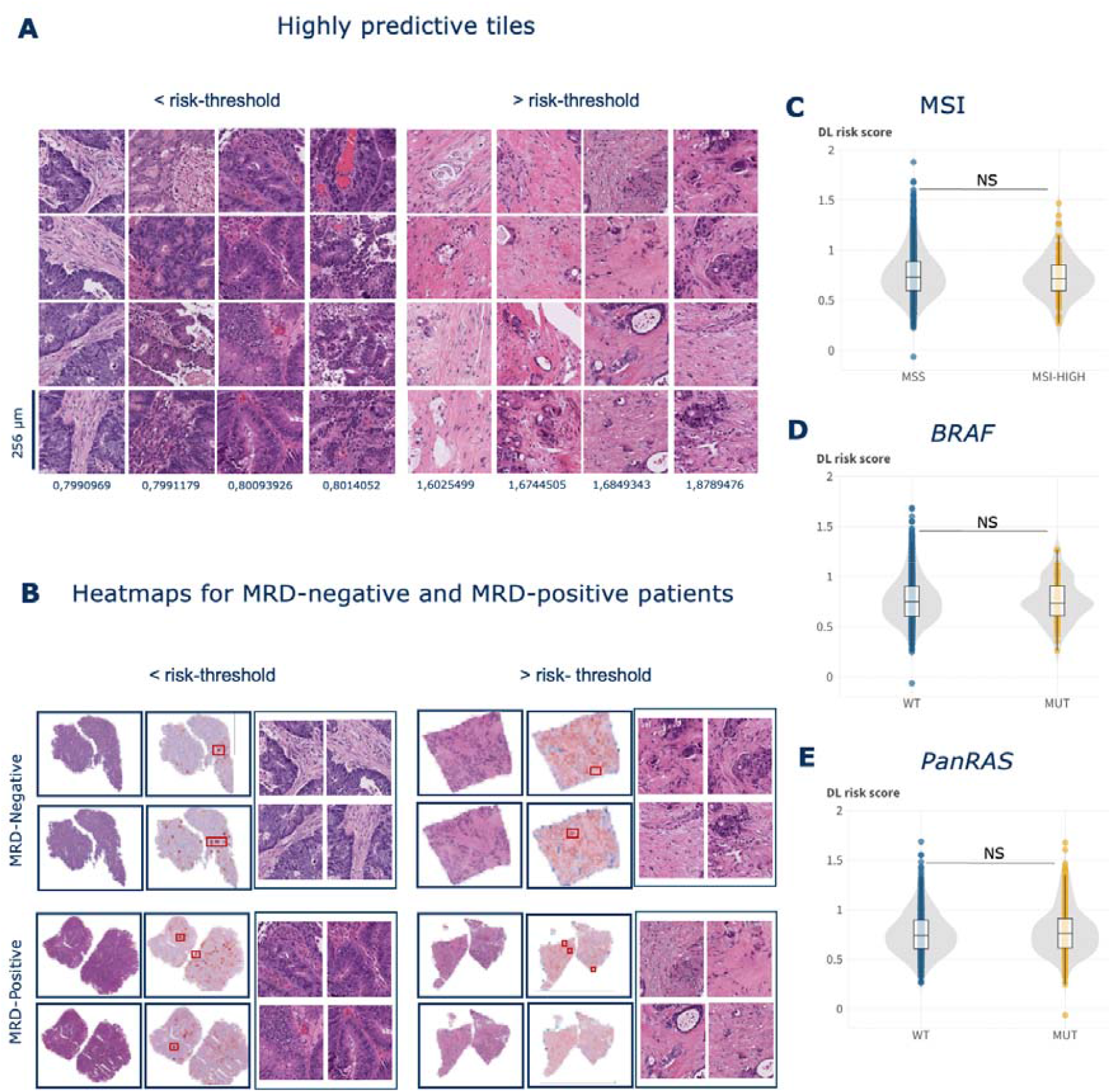
Morphological and molecular features of the DL risk score. (A) Highly predictive tiles for patients below the DL risk-threshold and above the DL risk-threshold exemplarily with DL score reported. (B) Whole slide patient heatmaps showing the DL prediction score, red indicating high-risk, and blue indicating low-risk. Box plot showing distribution of DL risk score among (C) MSI status (D) *BRAF-V600E* mutational status and (E) *PanRAS* mutational status. *P*-Value calculated using Kruskal-Wallis test. Figure was created using Flourish (https://flourish.studio/). DL= Deep Learning, MRD= molecular residual disease, WT= wildtype, MUT=mutation, MSS=microsatellite stable, MSI= microsatellite instability, NS=not significant

In the DL low-risk classified patients, the morphological analysis revealed a variety of benign histopathological tissue features (Supplementary Figure 3A). As the DL score increased, the histological image tiles still below the risk threshold displayed moderately differentiated tumor components. These samples still displayed a balanced tumor-stroma ratio and tumor glands with tubular to cribriform architecture, indicating an intermediate phenotype between DL low and DL high-risk morphological characteristics (Figure 3A-B). The images, above the risk threshold, displayed high-grade tumor cells with a significant desmoplastic stroma reaction. There was a high a intratumoral stroma fraction, and the presence of tumor buds/poorly differentiated clusters, which are known to be associated with a higher recurrence risk (Figure 3A-B).^32–36^ Taken together, we observed a clear morphological continuum mirroring the progression from DL low to DL high-risk tumors. Moreover, we analysed the distribution of the DL risk score with clinically relevant molecular information namely MSI status, *BRAF* and *RAS* mutational status (Figure 3 C-F). We found that the distribution was very similar for all these factors, suggesting that our DL model independently detects and accounts for additional prognostically relevant morphological features.

In summary, although our histopathology DL model was trained without human annotations, and solely on non-processed WSIs, we found that the model learned to pay attention to regions linked to tumor biological features plausibly associated with prognosis, thereby synergizing with ctDNA. Moreover, our findings are consistent with previous DL-based end-to-end prognostication approaches in CRC based on H&E histopathology alone^22,37^.

## Discussion

CRC can often be cured through surgery, but a subset of patients experience relapse, which is associated with high mortality. To mitigate this risk, ACT is administered to locally advanced CRC patients post-surgery. However, the majority of these patients do not benefit from such treatment, which is associated with substantial side effects^38^. Decades of research have focused on identifying potential biomarkers to administer ACT selectively to high-risk individuals who would benefit the most, while withholding it from low-risk individuals. To date, one of the most promising biomarkers for this purpose is ctDNA. Measurement of MRD through ctDNA is non-invasive, robust, and highly prognostic. However, ctDNA does not capture the tumor’s interaction with its microenvironment—the complex spatial ecology of tumors^39^-nor the tumor morphology itself. This is a limitation of ctDNA as a biomarker given that, in addition to conventional histopathology tumor features, the interplay between tumors and their microenvironment has been demonstrated to be highly prognostic and predictive over the years. In our study, we demonstrate that combining DL-based risk assessment with MRD measurement further enhances prognostic capabilities: MRD-negative patients who were predicted to be at high-risk for relapse by our DL model had a significantly longer DFS if treated with ACT, whereas in MRD-negative patients with a DL-based low-risk status no DFS benefit was seen for those receiving ACT (Figure 2F). These observations suggest that healthcare providers may identify a subset of patients who are at risk for relapse but are not detected through current diagnostic tools, including a diagnostic as innovative as ctDNA. Previous studies developing DL-based prognostication systems failed to provide evidence for potentially different chemotherapy efficacy across DL categories, by which all potential therapeutic implications of these models remain speculative^23,40^

To our knowledge, our study provides the first evidence suggesting that a DL risk assessment algorithm may indicate therapy efficacy in a real world setting in CRC. This combined approach may improve patient selection, suggesting a way how ACT could be restricted to those patients who are most likely to benefit from it. Furthermore, our DL method is using the latest state-of-the-art models, is fully open source and can be reused and adapted by anyone.

### Limitations

A limitation of our study is that integrating our insights into clinical routine requires further evaluation in additional cohorts, ideally in a prospective manner. Despite this, our study, encompassing thousands of patients across different ethnicities, represents one of the largest studies in this field. Moreover, we utilized a state-of-the-art foundation model for digital pathology analysis, UNI^30^. This is particularly relevant for clinical translatability, as the capabilities of foundation models are rapidly advancing, suggesting that further performance gains are conceivable with improved DL models. Nevertheless, medical device approval in Japan, the US, and the European Union requires a static piece of software that cannot be easily updated. Therefore, like any other DL-based biomarker, our method may be outdated by the time of clinical approval. We urge regulators and policymakers to work towards enabling the update of DL-based biomarkers with the latest technologies.

## Conclusion

Despite these limitations, our data show that the excellent prognostic performance of ctDNA in CRC can be further improved by DL-based end-to-end assessment of routine pathology slides. After prospective validation, this approach provides a plausible and comprehensive strategy for relapse risk assessment with potential therapeutic implications.

## Data Availability

https://github.com/mahmoodlab/uni

https://github.com/KatherLab/marugoto/tree/survival-transformer/marugoto/survival

## Ethics statement

The experiments in this study were carried out according to the Declaration of Helsinki and the International Ethical Guidelines for Biomedical Research Involving Human Subjects by the Council for International Organizations of Medical Sciences (CIOMS). The present study also adheres to the “Transparent reporting of a multivariable prediction model for individual prognosis or diagnosis” (TRIPOD) statement.20. The Ethics Board at the Medical Faculty of Technical University Dresden (BO-EK-444102022) and Institutional Review Board of the National Cancer Center Japan (2023-207) approved of the overall analysis in this study. The patient sample collection in each cohort was separately approved by the respective institutional ethics board.

## Author contributions

CMLL, HB, JNK and TY conceptualised the study. HD, TN, TM, SM, DK, HT, IT, TK, EO provided clinical and scanned whole slide image data for the GALAXY cohort. TY, DW, MH, HB provided clinical and scanned whole slide image data for the DACHS cohort. CMLL curated the source data. SS, XJ, MvT implemented the deep learning algorithm. SS developed the code for data analysis and visualisation. CMLL and SS planned and conducted the experiments. CMLL interpreted the data. HSM, ZIC, JNK assisted with the interpretation of results. NR and SF did the pathological interpretation of the results. CMLL wrote the first draft of the manuscript. All authors revised the manuscript draft, contributed to the interpretation of the data and agreed to the submission of this paper.

## Supplementary Figures and Tables

**Supplementary Figure 1:**
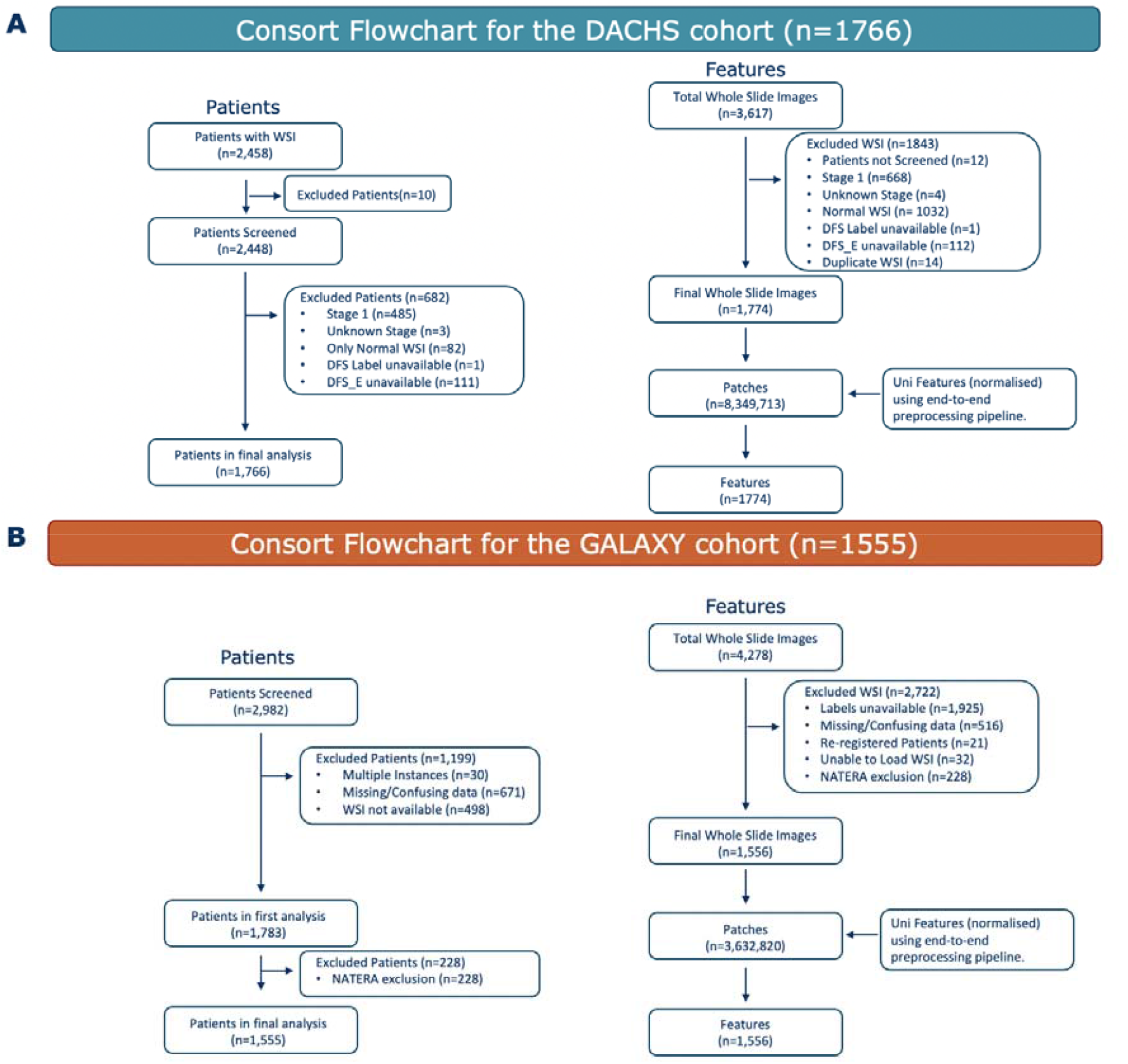
Consort diagram for both cohorts. Flowchart showing initial screened patients and WSIs and exclusion criterias for (A) the DACHS cohort and (B) the GALAXY cohort. WSIs= whole slide images, DACHS=Darmkrebs: Chancen der Verhütung durch Screening Study

**Supplementary Figure 2:**
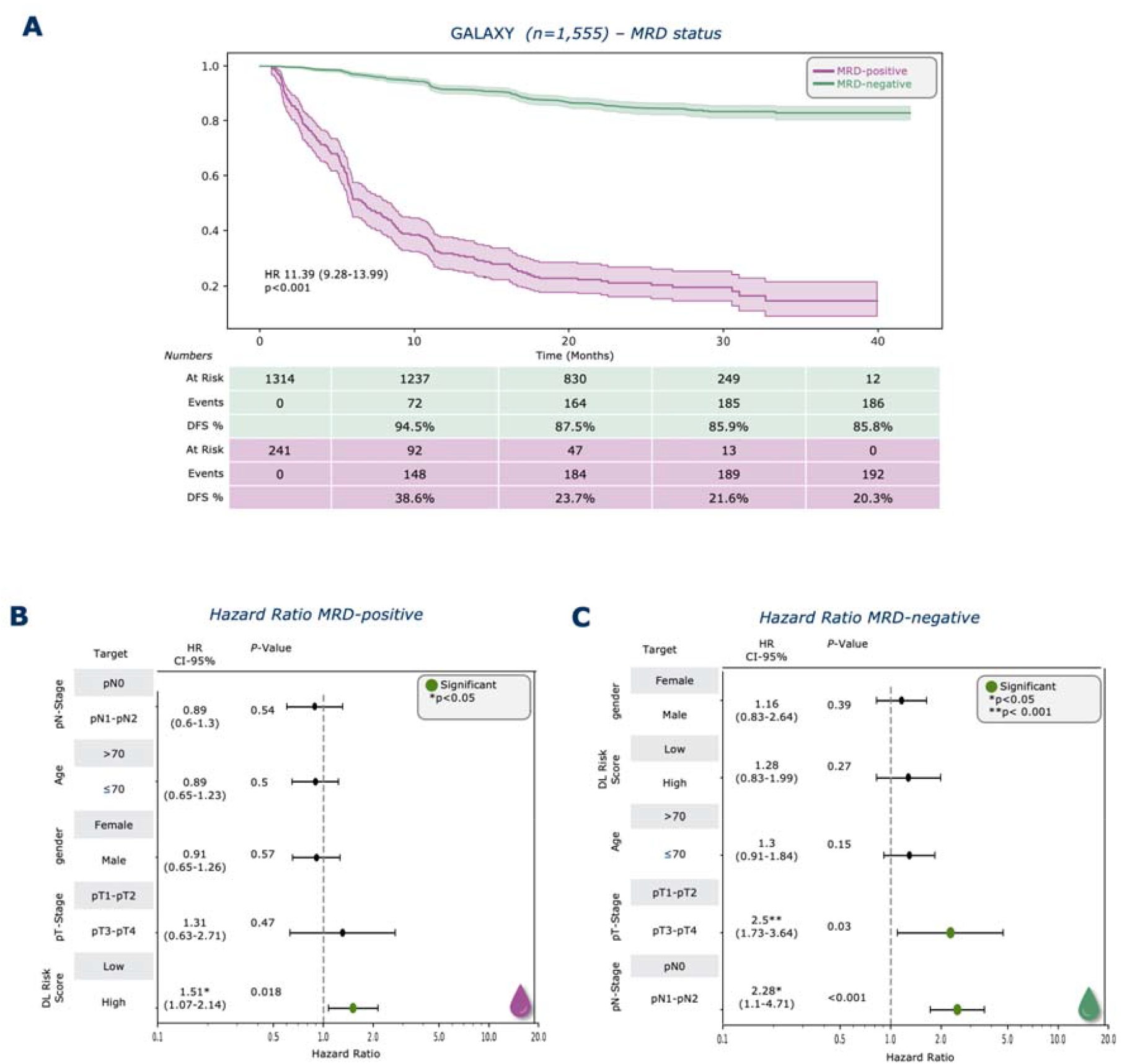
MRD status is predictive of survival outcomes and Multivariate analysis for MRD-subgroups. (A) Kaplan-Meier curves for DFS stratified by MRD-positive and MRD-negative patients. Forest plot showing multivariate cox regression analysis for (B) MRD-positive and (C) MRD-negative subgroup including the covariates gender, age, DL risk score, pathological Nodal Stage (pN-Stage), pathological Tumor Stage (pT-Stage) and their association with DFS. HR and 95% CI were calculated by the Cox proportional hazard model. *P*-value was calculated using the two-sided log-rank test (*p<0.05, ** p<0.001). Plot were generated using lifelines package in Python 3.11.5 DFS=disease-free survival, DL=Deep Learning, MRD=molecular residual disease, HR=Hazard ratio, CI=Confidence interval.

**Supplementary Figure 3:**
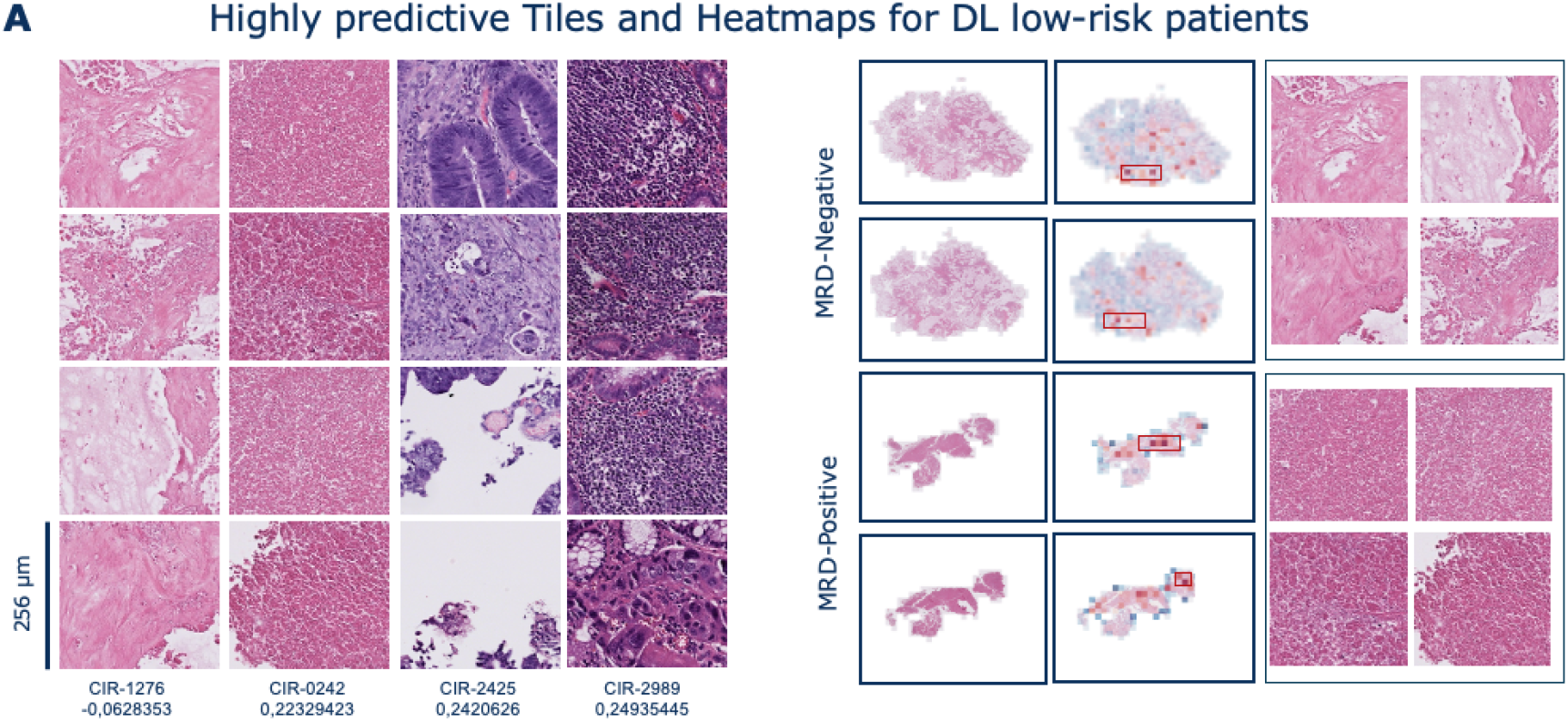
Morphological and molecular features for DL low-risk score. (A) Highly predictive tiles and Whole slide heatmaps for patients with lowest DL risk score. Red indicating high-risk, and blue indicating low-risk. DL= Deep Learning, MRD= molecular residual disease.

## Declaration of generative AI and AI-assisted technologies in the writing process

During the preparation of this study the author(s) used GPT-4 for grammar and spelling checking. After using this tool/service, the author(s) reviewed and edited the content as needed and take(s) full responsibility for the content of the publication.

